# Statistical framework for studying the spatial architecture of the tumor immune microenvironment

**DOI:** 10.1101/2021.04.27.21256104

**Authors:** Christopher Wilson, Ram Thapa, Jordan Creed, Jonathan Nguyen, Carlos Moran Segura, Travis Gerke, Joellen Schildkraut, Lauren Peres, Brooke L. Fridley

**Author notes:** Correspondence to: Brooke L. Fridley, Ph.D., 12902 Magnolia Drive, Moffitt Cancer Center, Tampa, FL 33612. None of the authors have conflict-of-interest or financial disclosures to declare. Email Addresses.

## Abstract

New technologies, such as multiplex immunofluorescence microscopy (mIF), are being developed and used for the assessment and visualization of the tumor immune microenvironment (TIME). These assays produce not only an estimate of the abundance of immune cells in the TIME, but also their spatial locations; however, there are currently few approaches to analyze the spatial context of the TIME. Thus, we have developed a framework for the spatial analysis of the TIME using Ripley’s *K*, coupled with a permutation-based framework to estimate and measure the departure from complete spatial randomness (CSR) as a measure of the interactions between immune cells. This approach was then applied to ovarian cancer using mIF collected on intra-tumoral regions of interest (ROIs) and tissue microarrays (TMAs) from 158 high-grade serous ovarian carcinoma patients in the African American Cancer Epidemiology Study (AACES) (94 subjects on TMAs resulting in 259 tissue cores; 91 subjects with 254 ROIs). Cox proportional hazard models were constructed to determine the association of abundance and spatial clustering of tumor-infiltrating lymphocytes, cytotoxic T-cells, and regulatory T-cells, and overall survival. We found that EOC patients with high abundance and low spatial clustering of tumor-infiltrating lymphocytes and cytotoxic T-cells in their tumors had the best overall survival. In contrast, patients with low levels of regulatory T-cells but with a high level of spatial clustering (compare to those with a low level of spatial clustering) had better survival. These findings underscore the prognostic importance of evaluating not only immune cell abundance but also the spatial contexture of the immune cells in the TIME. In conclusion, the application of this spatial analysis framework to the study of the TIME could lead to the identification of immune content and spatial architecture that could aid in the determination of patients that are likely to respond to immunotherapies.

## 1. Introduction

Immunology has been a break-through area in the treatment of cancer(2, 3). One of the most important findings is the use of agents to block immune checkpoints to activate antitumor immunity. Immune checkpoints are the mechanism by which the immune system maintains self-tolerance. That is, immune checkpoints are regulators of the immune system and prevent the immune system from attacking “good” cells. In the case of cancer, the cancerous cells hijack this mechanism to protect themselves from being attacked by the immune system(4). The use of checkpoint inhibitors in the treatment of cancer has been a revolutionary approach and has resulted in the development of numerous checkpoint inhibitors, such as CTLA-4 and PD-1/PD-L1 inhibitors.

Tumors with a dense infiltrate of lymphocytes, also known as tumor-infiltrating lymphocytes (TILs), are consistently associated with more favorable outcomes among cancer patients(5-7). However, abundance alone may not explain a patient’s clinical outcome, and consideration of the spatial architecture of the immune tumor immune microenvironment (TIME) may shed new light on clinical outcomes and response to immunotherapies. Lee, et al. showed that diffuse large B cell lymphoma tumors with similar densities of TILs had heterogeneous spatial patterns of cytotoxic T-cells(8), and a study in colorectal cancer observed that cell-to-cell distances and spatial heterogeneity were more promising as prognostic biomarkers than cell densities(9).

Many technologies have been developed to study the TIME. One study approach is the use of multiplex immunofluorescence (mIF) microscopy which provides both a summary of the number of cells positive for a given immune marker (e.g., abundance or density) but also the spatial locations of the positive cells. By having spatial locations of the cells positive for the various immune markers, one can determine spatial clustering and co-occurrenceof immune cells. mIF can be applied to both regions of interest (ROI) selected from a stained tumor slide or tissue microarrays (TMAs)(10). Many challenges arise with the use of data resulting from TMAs. In particular, the tissue area can become folded or ripped due to the “slicing” for different experiments, leading to imperfections in the shape and the ability to measure all cells in the area. These imperfections can lead to TMAs that have sections where no cells exist, as depicted in **Figure 1A**. In contrast, ROIs typically do not exhibit this artifact in the data acquisition (**Figure 1B**).

**Figure 1:**
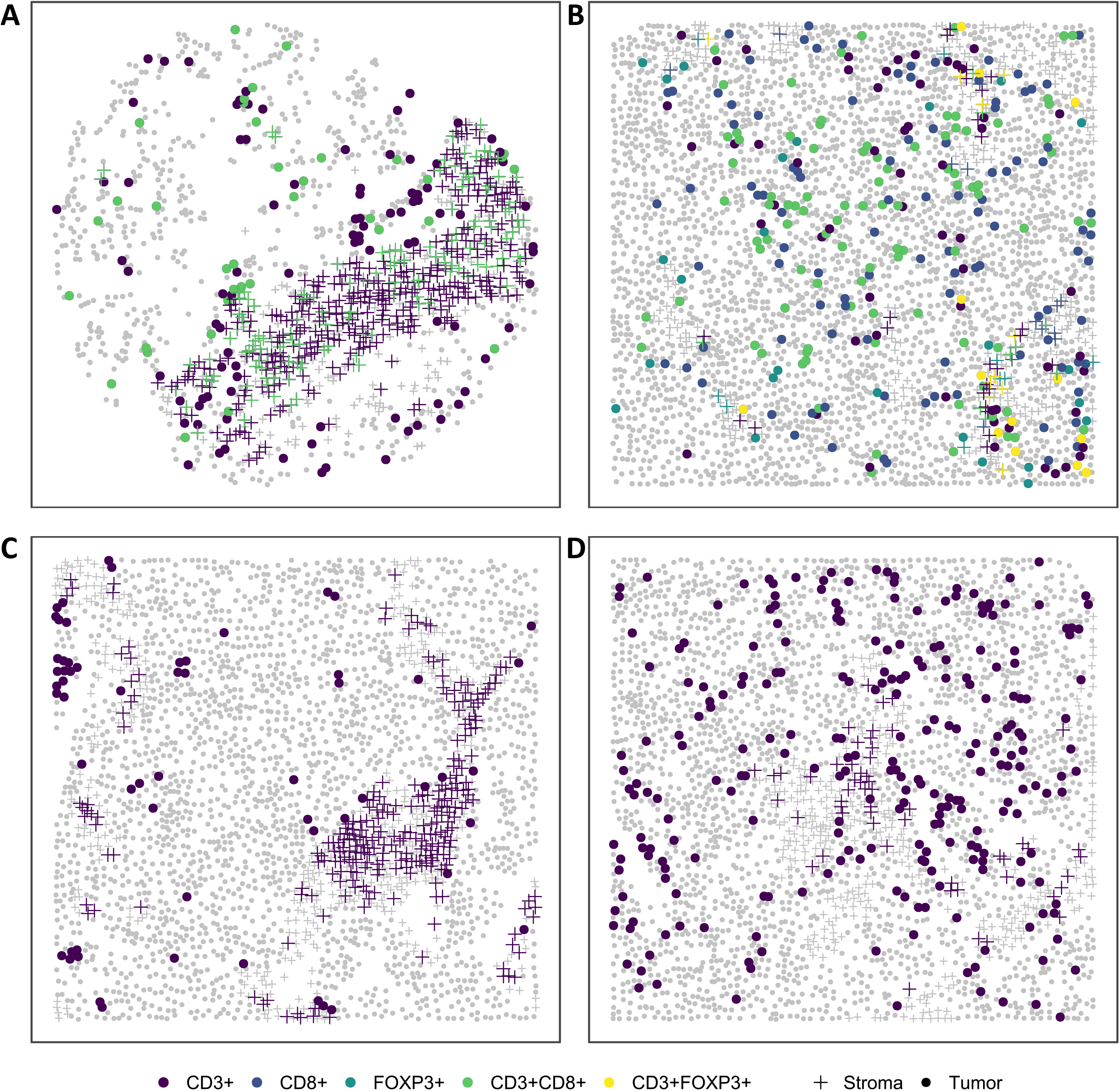
Example of IF data from **(A)** a TMA core sample and **(B)** an intra-tumoral region of interest (ROI). As illustrated in the two example figures, TMAs tend to have more “holes” and uneven cell density as compared to ROIs. Examples of clustering patterns observed in the intra-tumoral ROIs; **(C)** location of the CD3+ cells in which the pattern is close to complete spatial randomness (CSR); and **(D)** location of CD3+ cells in which the pattern deviates from CSR.

The most common analysis of data from the TIME involves the use of the summary measures representing immune content in the entire sample (i.e., % of CD3+ cells, density). However, this type of analysis ignores the spatial architecture of the immune cells within the tumor, which can vary between tumors. As illustrated in **Figure 1**, some tumors show clustering of TILs (CD3+ cells; **Figure 1C**), while other tumors show more dispersion of TILs **(Figure 1D)**. While there have been studies that attempt to describe the relationship between spatial clustering of immune cells and patient outcomes using such measures as nearest-neighbor distance (NND)(11), Hypothesized Interaction Distribution (HID) (12, 13), Morisita-Horn index(14), and Mander’s correlation coefficient(15), many approaches fail to account for issues related to: correlation between spatial and abundance measures of immune cells; edge/border effects; normalization of measures across samples, and regions in which no cells were able to be measured(16, 17). Thus, we have developed a framework for the spatial analysis of the TIME using Ripley’s *K*(1), coupled with a permutation-based framework to estimate and measure the departure from complete spatial randomness (CSR) as a measure of the relationships and interactions between immune cells. We applied this analysis framework to epithelial ovarian cancer (EOC), the deadliest gynecologic malignancy in the U.S.(18). Specifically, we will characterize the TIME and explore links between immune cell abundance and their spatial characteristics, and overall survival (OS) among EOC patients.

## 2. Methods and Materials

### 2.1 Study Population and Immunofluorescence Assays

The African American Cancer Epidemiology Study (AACES) is a population-based case-control study of 595 African American (AA) women with EOC residing in 11 geographic locations in the U.S. and 752 controls enrolled between December 2010 and August 2016(19). Cases were identified through cancer registries and hospitals and were eligible for the study if they were aged 20-79 years, self-reported AA race, and resided in one of the 11 geographic locations. Study participants completed a telephone survey at baseline, and for ∼90% of the cases, formalin-fixed paraffin-embedded (FFPE) tissue blocks of the primary tumor were procured. For 75% of cases with tissue, twenty-five sections were cut from each tissue block, and TMAs were created for the other 25%. A centralized pathology review was completed to confirm diagnosis and histology. A systematic collection of vital status and follow-up data through linkages with cancer registries and the National Death Index (NDI) has been conducted annually. All participants provided verbal consent at the time of the baseline telephone interview, and written informed consent was obtained for the procurement of tissue specimens and collection of medical records.

As the distribution of immune cells in the tumor microenvironment and their association with outcomes differ according to histotype (20), we focused on the most common and one of the deadliest histotypes, high-grade serous carcinoma (HGSC)(21), to limit contributions of disease heterogeneity. To determine immune profiles of these tumors, mIF staining was completed using the Opal™ chemistry and multispectral microscopy Vectra system (Akoya Biosciences). Tumors were stained for one panel including seven fluorophore-labeled markers: CD3, CD8, FOXP3, CD11b, CD15, DAPI and pancytokeratin (PanCK). After staining, slides were scanned, and image capture was performed with the Vectra®3 Automated Quantitative Pathology Imaging System (Akoya Biosciences) with images are exported from InForm (Akoya Biosciences) and loaded into HALO (Indica Labs, New Mexico) for quantitative image analysis. Coordinates of the cell locations are based on pixels of the image where the image resolution is 0.4977 microns per pixel (Mpp). For these analyses, we focused on tumor-infiltrating lymphocytes (CD3+) and relevant T-cell subsets (CD3+CD8+ cytotoxic T-cells; CD3+FOXP3+ regulatory T-cells). In addition to TMAs, mIF was performed on whole slide images, where three ROIs from the intratumoral region of each tissue section were selected for analysis (e.g., 100% tumor cells by cellularity and PCK expression). A summary of the study participants is presented in **Supplemental Table 1**, where 94 subjects were on TMAs (259 samples) and 91 subjects had ROIs (254 samples), with 27 subjects with both TMA and ROI data.

**Table 1:**
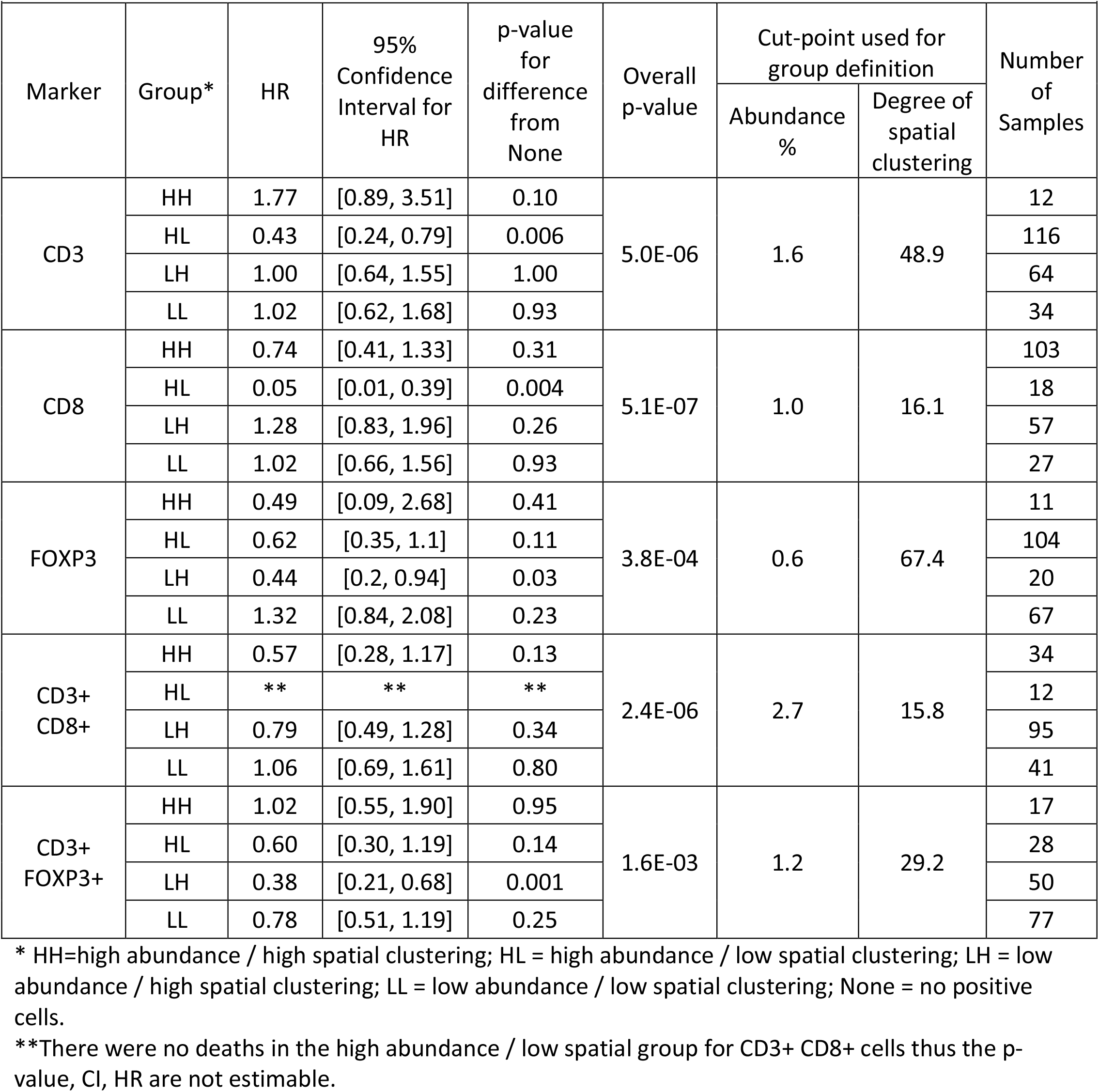
Results from survival analysis of the immune marker abundance and spatial clustering from the ROIs involving 91 subjects (254 ROIs). Degree of spatial clustering based on permutation-based estimate of CSR. Models were adjusted for age at diagnosis and stage. Group None is reference group.

| Marker | Group* | HR | 95% Confidence Interval for HR | p-value for difference from None | Overall p-value | Cut-point used for group definition |  | Number of Samples |
| --- | --- | --- | --- | --- | --- | --- | --- | --- |
|  |  |  |  |  |  | Abundance % | Degree of spatial clustering |  |
| CD3 | HH | 1.77 | [0.89, 3.51] | 0.10 | 5.0E-06 | 1.6 | 48.9 | 12 |
|  | HL | 0.43 | [0.24, 0.79] | 0.006 |  |  |  | 116 |
|  | LH | 1.00 | [0.64, 1.55] | 1.00 |  |  |  | 64 |
|  | LL | 1.02 | [0.62, 1.68] | 0.93 |  |  |  | 34 |
| CD8 | HH | 0.74 | [0.41, 1.33] | 0.31 | 5.1E-07 | 1.0 | 16.1 | 103 |
|  | HL | 0.05 | [0.01, 0.39] | 0.004 |  |  |  | 18 |
|  | LH | 1.28 | [0.83, 1.96] | 0.26 |  |  |  | 57 |
|  | LL | 1.02 | [0.66, 1.56] | 0.93 |  |  |  | 27 |
| FOXP3 | HH | 0.49 | [0.09, 2.68] | 0.41 | 3.8E-04 | 0.6 | 67.4 | 11 |
|  | HL | 0.62 | [0.35, 1.1] | 0.11 |  |  |  | 104 |
|  | LH | 0.44 | [0.2, 0.94] | 0.03 |  |  |  | 20 |
|  | LL | 1.32 | [0.84, 2.08] | 0.23 |  |  |  | 67 |
| CD3+ CD8+ | HH | 0.57 | [0.28, 1.17] | 0.13 | 2.4E-06 | 2.7 | 15.8 | 34 |
|  | HL | ** | ** | ** |  |  |  | 12 |
|  | LH | 0.79 | [0.49, 1.28] | 0.34 |  |  |  | 95 |
|  | LL | 1.06 | [0.69, 1.61] | 0.80 |  |  |  | 41 |
| CD3+ FOXP3+ | HH | 1.02 | [0.55, 1.90] | 0.95 | 1.6E-03 | 1.2 | 29.2 | 17 |
|  | HL | 0.60 | [0.30, 1.19] | 0.14 |  |  |  | 28 |
|  | LH | 0.38 | [0.21, 0.68] | 0.001 |  |  |  | 50 |
|  | LL | 0.78 | [0.51, 1.19] | 0.25 |  |  |  | 77 |

\* HH=high abundance / high spatial clustering; HL = high abundance / low spatial clustering; LH = low abundance / high spatial clustering; LL = low abundance / low spatial clustering; None = no positive cells.
\*\*There were no deaths in the high abundance / low spatial group for CD3+ CD8+ cells thus the p-value, CI, HR are not estimable.

### 2.2 Ripley’s *K* and Complete Spatial Randomness

In our proposed framework, the locations of the immune positive cells in the TIME can be thought of as a spatial point process. The arrangement of these cells may not follow the assumption of complete spatial randomness (CSR) for homogenous spatial processes, where positive cells occur at the same rate λ for the entire region. Attraction (clustering), repulsion, and competition (dispersion) are all examples of interactions that would lead to a violation of CSR. Ripley’s *K*(1) and Besag’s L(22) are popular measures to quantify these interactions by counting the number of neighboring cells within a radius for each positive immune cell, normalizing by the maximum number of pairwise distances, and correcting for border effects. **Figure** Error! Reference source not found.**1C** shows an ROI that exhibits a spatial process that is close to CSR, while **Figure 1D** shows an ROI that violates the CSR assumption.

A common measure to assess the CSR is Ripley’s *K*, which is computed at several rings with varying radii, r, to help understand how the point process changes with scale. The sample statistic for this quantity is estimated by:

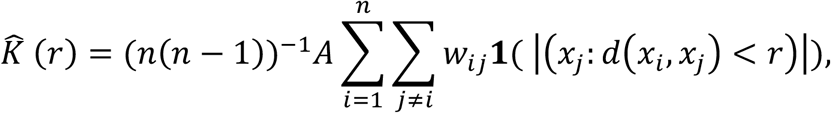

Where *n* is the number of cells, A represents the area of the region of interest, *W*_*ij*_ corresponds to the edge correction, and 1 |*x*_*j*_: *d*(*x*_*i*_, *x*_*j*_) < r | is an indicates whether the *j*^th^ is a neighbor of the *i*^th^ cell (the Euclidean distance between cells i and j is less than r), where the expected value for 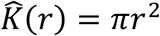 The edge correction accounts for undercounting of the number of neighboring cells when a cell on the periphery. The difference between the observed and expected values, 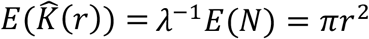 where E(N) is the expected number of cells and λis the intensity of the cell, helps determine the degree of regularity or clustering. A positive difference corresponds to a higher degree of clustering than expected, while a negative difference is evidence of a regular pattern existing(1, 23, 24). Besag proposed the following modification to Ripley’s *K*, 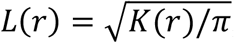 where the expected value is r An advantage of analyzing L(r) is that the expected value is grows linearly with r as opposed to quadratically, additionally the square root is a commonly used variance stabilizing transformation(22). Using simulated data, **Supplemental Figure 1** illustrates the connection between the type of spatial arrangement of immune cells (**Supplemental Figure 1A**) (i.e., clustered or attraction, spatial randomness (CSR), or regularized or repulsion) and the degree of spatial clustering measured as *L(r) – r* (**Supplemental Figure 1B**)

Both Ripley’s *K* and Besag’s *L* can be calculated in a univariate form as described above, or a bivariate form where you could explore co-occurrence of types of immune cells, for example, the spatial pattern of cytotoxic T-cells around regulatory T-cells. A general formula is given by

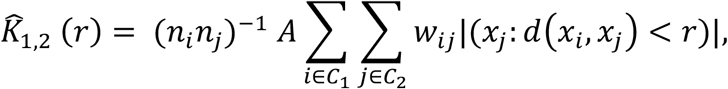

*C*_1_ and *C*_2_ are the set of cells that are determined to be in type 1 or 2, respectively. The expected value of 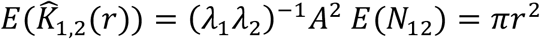, where E(N_12_) is the expected number of cells of type 2 within distance r of a cell of type 1, and λ_1_ and λ_2_ are the intensity functions for cell type 1 and 2, respectively. Similar adjustments have been proposed to linearize to aid in interpretation(25).

Ideally, these point processes occur in a rectangular or circular region of space, however, the region where cells appear are not necessarily perfect rectangles or circles. To that end, we can still estimate Ripley’s *K* over the convex hull of the cells measured in the sample. The convex hull is the smallest convex set that contains every cell and line segment connecting any to cells. The “edge effect” results from cells on the periphery of the tissue sample lose neighboring cells that are located outside the sampled region, also needs to be accounted for in the computation of Ripley’s *K*. Two common edge corrections are called isotropic and translation(26, 27). The translation edge correction is conducted by sequentially shifting the image by the distance between a cell and an adjacent cell where the weight depends on the amount of area that is in the intersection of the two images(23). An isotropic edge correction, for circular or rectangular regions, weights each pair of points based on how much of the circumference of a circle centered around one point and going through the other is outside of the region of interest. The translation and isotropic edge correction methods can accommodate settings when there are a small number of positive cells in the tissue samples(23). Additionally, little difference was observed in the estimate of Ripley’s *K* using these two border correction methods to IF data, with a correlation value around 1.0 (data not shown).

As many of the methods for spatial point processes were developed for the analysis of one dataset, normalization of the spatial measurements across multiple samples is needed. In the proposed framework, normalization is completed by comparing the observed value of Ripley’s *K* to the estimate of Ripley’s *K* under CSR. Unlike ROIs selected from whole tissue sections, TMAs have many cores with regions that are folded or torn; in these cases, it appears that there are no cells present at various locations. This will result in the appearance of the intensity function of the observed point process to not be constant across the entire region, while the true underlying process may have been homogenous. Hence, use of the theoretical expected value of *K* and *L* under CSR would not be appropriate. To overcome this challenge, a permutation approach is used to obtain a robust estimate of these spatial statistics under CSR.

### 2.3 Permutation-based measure of CSR

Non-parametric statistical methods are commonly used when models’ assumptions are not met. A class of non-parametric methods are those based on permutation or Monte-Carlo methods, where the sampling distribution of interest under the null hypothesis (i.e., CSR) is estimated by randomly assigning the labels and computing the desired statistic. This process is carried out many times and the resulting distribution of the statistic is an empirically derived distribution. In this context, each cell is labeled based on whether a certain marker is present or absent. Next, these labels are randomly assigned to each cell and the measure of spatial clustering is computed, maintaining the number of cells present and absent of a marker. This process is repeated a larger number of times resulting in an empirically derived sampling distribution under the null hypothesis (i.e., CSR) for the computed spatial statistics (i.e., Ripley’s *K*). Using this empirical distribution, the mean is computed and used as the estimate of these spatial statistics under CSR(28).

### 2.4 Association of immune cell spatial clustering and survival from ovarian cancer

To measure the association of the level of spatial clustering of immune cells with clinical outcome, while also accounting for the abundance level, Cox proportional hazards models were used. Accounting for the abundance level is critical as computation of spatial measures require an adequate number of positive cells (i.e., negative relationship between abundance level and spatial clustering value, **Supplemental Figure 2**). Depending on the outcome or phenotype of interest, other statistical models could be used (i.e., linear models in the case of a continuous outcome).

**Figure 2:**
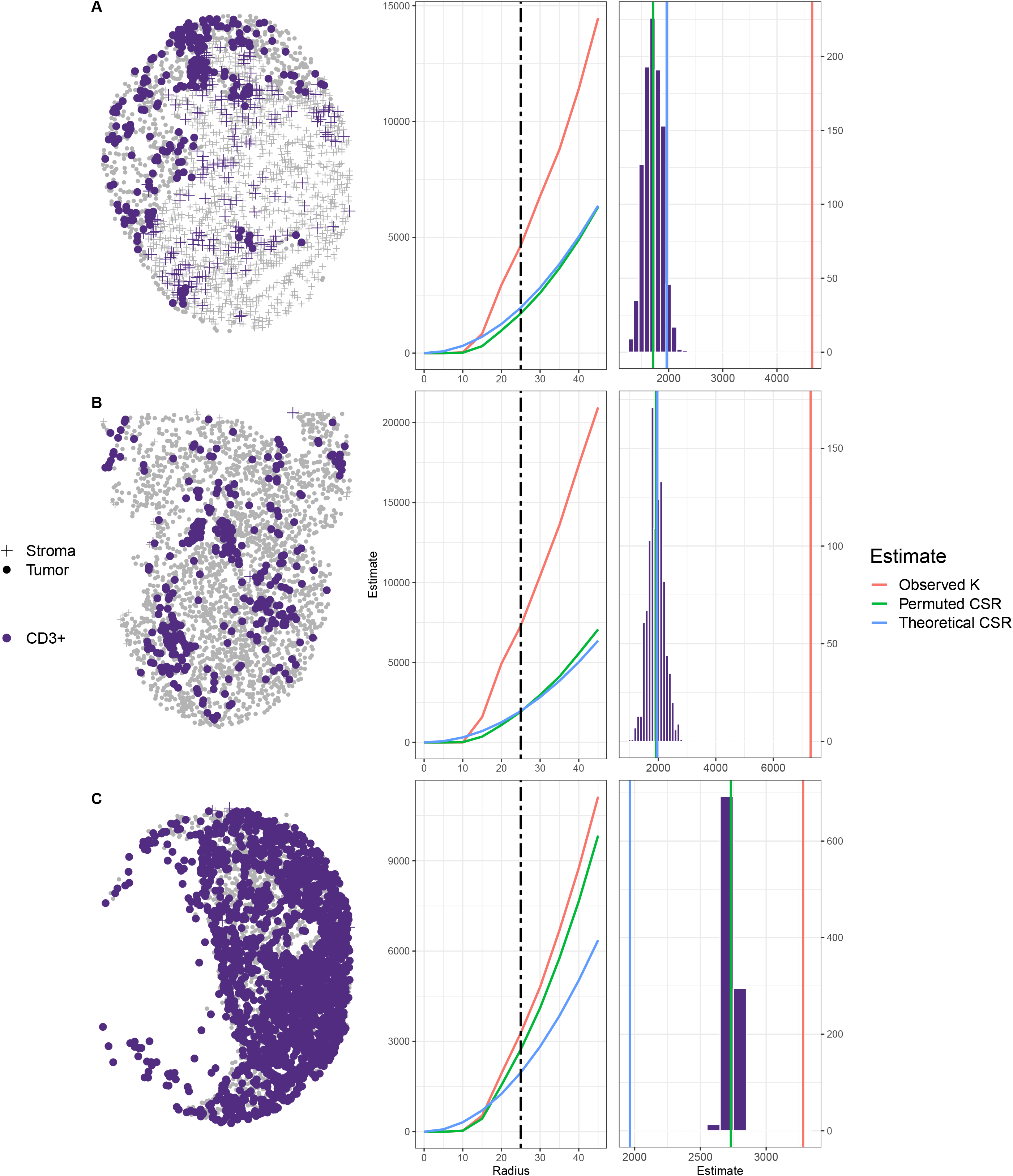
The empirical distribution of Ripley’s K under CSR generated by permutation with the translational edge correction for r = 50 and 100 for CD3+ cells. The observed value of Ripley’s K is represented by blue dotted line, the mean of the permutation-based CSR distribution is represented by the vertical black solid line, and the theoretical CSR computed assuming no “holes” in the tissue sample image is the vertical red line. As the level of missing cells increases (i.e., more “holes”) there is larger difference between the estimate of CSR based on theoretical computation using area and the estimate of CSR based on the mean of the empirically derived distribution.

In order to provide clinically interpretable results, the continuous measurement of abundance and spatial clustering were categorized (e.g., absent/present of immune marker, absent/low level/high level of abundance for immune marker). Typically, categorization takes place for each variable where values are assigned to groups depending on their relationship to the median, quartiles or some other threshold. Thus, an alternate for setting the pre-defined threshold is the “optimal cut-point”, which is selected to maximize the test statistic of interest (29, 30). From a statistical standpoint, the optimal cut-point is “data-snooping” since we are allowing the results from the statistical test to inform creation of categories of the variable(31). On the other hand, for discovery and hypothesis generation purposes, it is clinically useful to determine an optimal cut-point that can be validated in other studies with other technologies. Thus, for this study, we have chosen to use the “optimal” cut-point approach to determine thresholds that produce the largest difference in the survival curves.

A challenge in determining the optimal cut-points is that the number of samples in a group/category can get very small when categorizing across multiple variables. Thus, in the analysis to determine the association of immune cell abundance and spatial clustering, optimal cut-points were determined based on a grid search where the search was constrained to possible cut-points in which each group had at least 10% of samples (after removing samples with 0 abundance). This constrained approach ensured that each group had an adequate size for model stability. As Ripley’s *K* is not estimable in the case of no immune cells in a sample, this produced 5 groups: no immune cells; low abundance and low spatial clustering; low abundance and high spatial clustering; high abundance and low spatial clustering; and high abundance and high spatial clustering. The model that is reported is the model that had the largest statistical significance due to the abundance and degree of spatial clustering for each marker. The log-ratio test comparing the models was used to isolate the effect of the five-group stratification.

For the co-occurrence analysis involving two types of immune cells with OS, the bivariate version of Ripley’s *K* was used. As bivariate Ripley’s *K* is only estimable when both types of immune cells are present in a sample, the level of spatial clustering was only computed when both immune markers were present. Five categories were defined as AAN, APN, PAN, PPL, and PPH, where the first letter corresponds to the (A)bsence or (P)resence of the first cell type, the second letter represents the (A)bsence or (P)resence of the second cell type, and the last letter describes the level of spatial clustering (N)one/(L)ow/(H)igh. The marker was defined as present if at least one cell was positive for the immune marker in the sample. The threshold to determine low vs high spatial clustering based on the value of bivariate Ripley’s *K* led to the stratification which resulted in the smallest p-value from a log-ratio test comparing the models with just clinical information and a model with clinical information and the stratification based on abundance and spatial information. Note that the bivariate version of Ripley’s *K* is not a symmetric statistic. That is, analysis is first completed using the cytotoxic T-cell as the “anchor” or center for the computation of Ripley’s *K*, followed by the analysis using the regulatory T-cells as the “anchor”.

Using these pre-defined categories for the abundance and spatial clustering, association of each cell type and phenotype with OS following EOC was completed using Cox proportional hazards models for CD3+, CD8+, FOXP3+, CD3+CD8+, and CD3+FOXP3+. The model included clinical covariates of age at diagnosis and stage (I, II, III, IV) and accounted for the repeated measurements per tumor/subject. Analysis was completed separately for the intratumoral ROIs and TMAs. For the analysis of the TMAs, analyses were based only on the immune marker data in the tumoral compartment of each core in order to compare findings between the intratumoral ROIs and the TMAs (i.e., restricted analysis to the cells determined to be tumor based on PanCK marker and removed the cells within the stromal compartment). All statistical analyses were completed using RStudio with R version 4.0.4.

## 3 Results

### 3.1. Quality Control and Assessment

Prior to statistical analysis of the mIF data, quality assessment of the data was completed. In calling a cell as positive or negative for a marker, a machine learning algorithm within the HALO (Indica Labs, New Mexico) software was used where a threshold was determined based on the intensity measurements. As the sensitivity and specificity of this method is not 100%, there may be cases where a cell is classified for multiple phenotype combinations that are not possible, such as a cell being classified as both CD3+CD8+ (Cytotoxic T-cell) and CD3+FOXP3+ (regulatory T-cell or Treg) cell (**Supplemental Figure 3**). For this analysis, the locations for 1 – 87 cells on 133 out of 254 ROI samples and 1-73 cells on 63 out of 259 TMA cores **(Supplemental Figure 4**) were retained, but not the phenotype identification of cytotoxic T-cell and Treg. Instead, these cells were just considered to be positive for CD3+.

**Figure 3:**
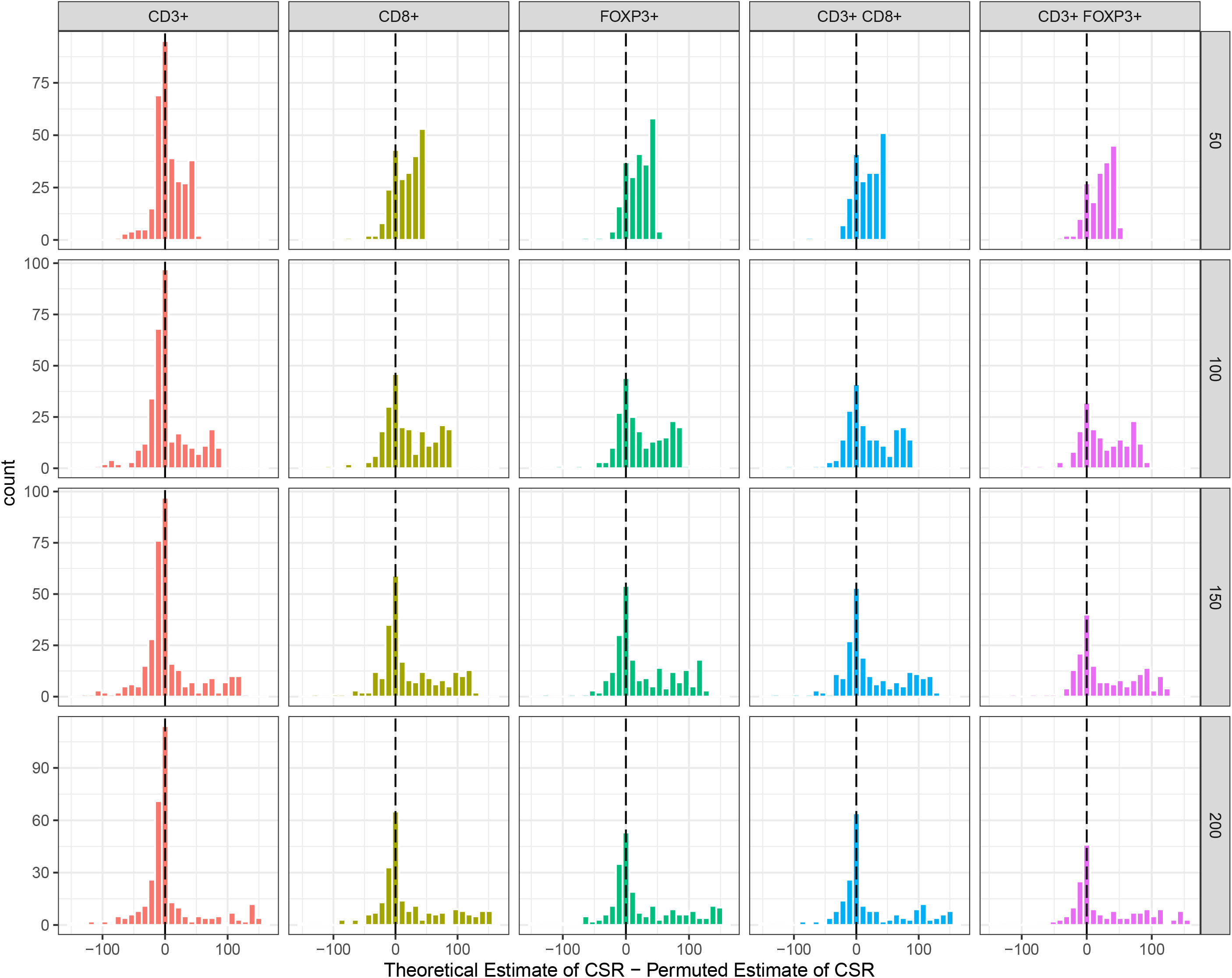
Histogram of the difference between the theoretical estimate and the permuted estimate of CSR for the 94 TMA samples for CD3+, CD8+, CD3+CD8+ (cytotoxic T-cell), FOXP3+ and CD3+FOXP3+ (Regulatory T-cell or Treg). The dashed black line represents the mean of distribution.

**Figure 4:**
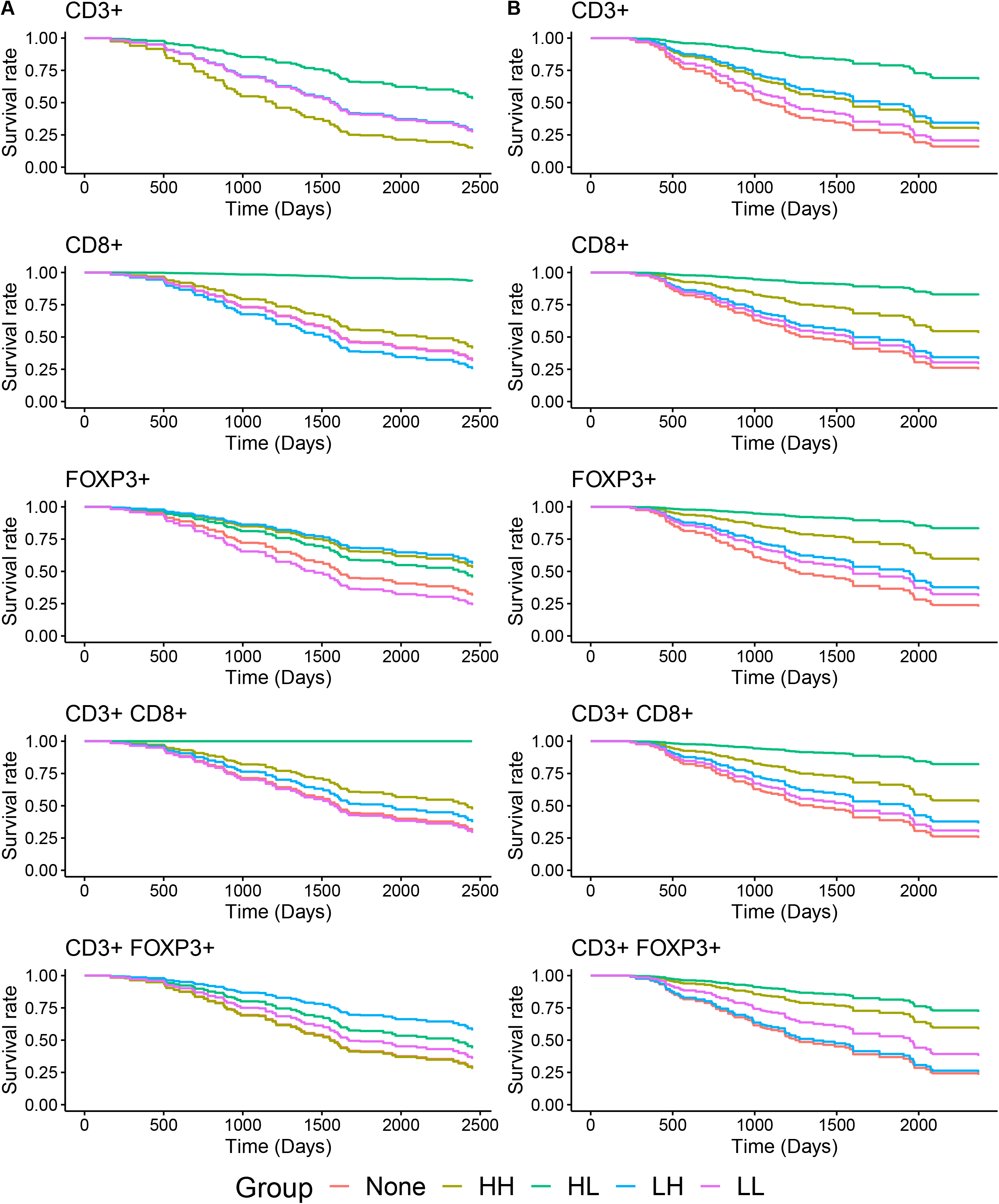
Predicted survival curves from Cox proportional hazard models for the CD3+, CD3+CD8+, and CD3+FOXP3+ cells where the degree of spatial clustering was based the permutation-based estimate of Ripley’s K under CSR (i.e., observed Ripley’s K – the mean of the empirical distribution of Ripley’s K under CSR); **(A)** results from intra-tumoral ROIs (91 subjects, 254 samples); **(B)** results from tumor compartment of TMAs (94 subjects, 259 samples). Models adjusted for age at diagnosis and stage within a repeated measures analysis framework.

### 3.2. Estimate of degree of spatial clustering and CSR in TMAs and ROIs

Following quality control, the levels of spatial clustering were estimated using the permutation-based value for CSR, where the degree of spatial clustering was computed as Ripley’s *K* – CSR permutation. **Figure 2** shows three TMA cores and the empirical sampling distribution of Ripley’s *K* under CSR for marker CD3. The first row (A) corresponds to a TMA core that does not have large areas where cells are absent, the second row (B) displays a TMA core with a moderate number of missing cells, and the third row (C) shows a TMA core with an extensive level of missing cells. As the level of missing cells increases, the difference between the theoretical and permutation-based estimates for Ripley’s *K* under CSR can increases. Similarly, the histogram of the distance between the permutation-based estimate of CSR and theoretical estimates of CSR is presented in **Figure 3 (TMA) and Supplemental Figure 5 (ROI)**. The mean of the difference (Theoretical estimate of CSR – Permuted estimate of CSR) is positive which indicates that the theoretical estimate of Ripley’s *K* under CSR is overestimated, resulting in a biased estimate of the degree of spatial clustering and incorrect association results with a phenotype of interest. In contrast, the Monte-Carlo estimate of Ripley’s *K* under CSR would provide a more accurate measure of spatial clustering which accounts for the unevenness in the cell distribution measured in the sample.

**Figure 5:**
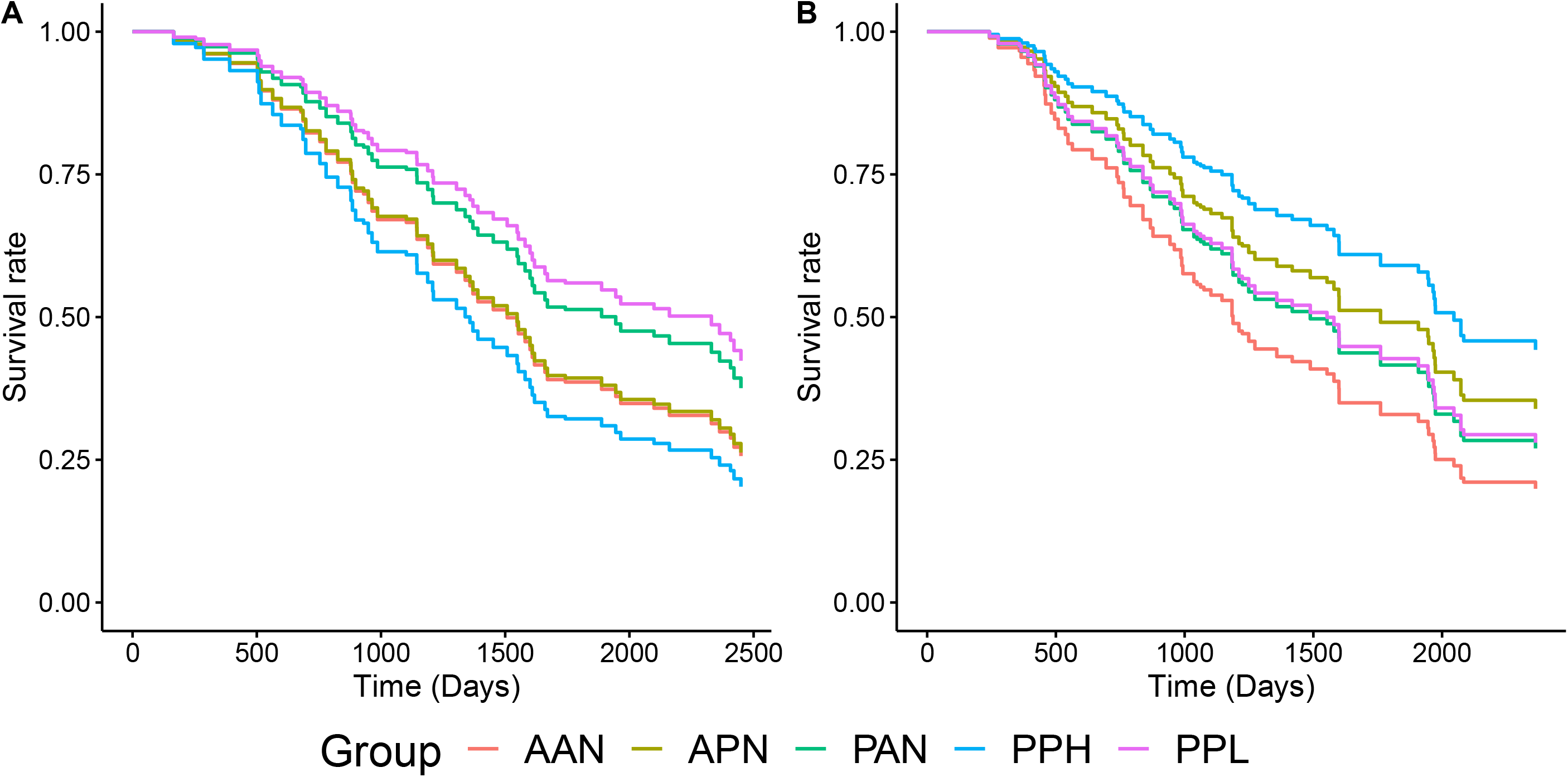
Predicted survival curves from Cox proportional hazard models for the degree of cooccurrence of CD3+ FOXP3+ and CD3+ CD8+(reference cell type) cells was based the permutation-based estimate of Ripley’s K under CSR (i.e., observed Ripley’s K – the mean of the empirical distribution of Ripley’s K under CSR); **(A)** results from intra-tumoral ROIs (91 subjects, 254 samples); **(B)** results from tumor compartment of TMAs (94 subjects, 259 samples). Models adjusted for age at diagnosis and stage within a repeated measures analysis framework.

### 3.3. Analysis of spatial clustering using univariate Ripley’s *K* and ovarian cancer survival

#### 3.3.1. Intratumoral ROIs

Cox proportional hazard models were fit to assess the association of the abundance and spatial clustering of cells positive for CD3+ (T-cells), CD8+, FOXP3+, CD3+CD8+ (cytotoxic T-cell), and CD3+FOXP3+ (regulatory T-cells), adjusting for age at diagnosis and stage, where the degree of spatial clustering was measured by the difference of the observed estimate of Ripley’s *K* from the permutation-based estimate of Ripley’s *K* under CSR. The estimates of abundance and univariate spatial clustering were collapsed into five categorial groups as described in **Section 2.4**. **Table 1** presents the hazard ratio (HR) estimates for the different groups with the “None” group representing the reference group. Survival curves for all five models are presented in **Figure 4A**. The optimal cut-point for determining high and low abundance was around 0.5 – 3% for the various cell types. Using the optimal cut-points for abundance (high vs low) and degree of spatial clustering (high vs low), there was evidence that EOC patients with high abundance and low spatial clustering (HL group) of CD3+, CD8+, and CD3+CD8+ cells in their tumors had the best OS. When restricting the analysis to those samples with high abundance based on the optimal cut-point, a significant difference was observed in the survival curves between patients with low and high spatial clustering of CD3+ (HR = 0.22, 95% CI (0.11, 0.47)), CD8+ (HR = 0.07, 95% CI (0.01, 0.54)), and CD3+CD8+ (HR and CI not estimable as no deaths in the high abundance / low spatial clustering group) immune cells, with EOC patients with low clustering having best survival (**Supplemental Table 2**).

**Table 2:** Results from survival analysis of the immune marker abundance and spatial clustering from the tumor compartment of the TMAs involving 94 subjects (259 cores on TMAs). Degree of spatial clustering based on permutation-based estimate of CSR. Models were adjusted for age at diagnosis and stage. Group None is reference group.

| Marker | Group* | HR | 95% Confidence Interval for HR | p-value for difference from “None” group. | Overall p-value | Cut-point used for group definition |  | Number of Samples |
| --- | --- | --- | --- | --- | --- | --- | --- | --- |
|  |  |  |  |  |  | Abundance % | Degree of spatial clustering |  |
| CD3 | HH | 0.55 | [0.25, 1.19] | 0.13 | 6.7E-12 | 6.0 | 22.4 | 26 |
|  | HL | 0.14 | [0.07, 0.30] | 0 |  |  |  | 38 |
|  | LH | 0.48 | [0.29, 0.78] | 0.003 |  |  |  | 111 |
|  | LL | 0.80 | [0.48, 1.32] | 0.38 |  |  |  | 129 |
| CD8 | HH | 0.39 | [0.12, 1.31] | 0.13 | 7.3E-08 | 5.3 | 22.1 | 9 |
|  | HL | 0.11 | [0.02, 0.50] | 0.004 |  |  |  | 17 |
|  | LH | 0.75 | [0.44, 1.28] | 0.29 |  |  |  | 67 |
|  | LL | 0.86 | [0.54, 1.36] | 0.52 |  |  |  | 69 |
| FOXP3 | HH | 0.30 | [0.08, 1.20] | 0.09 | 3.8E-07 | 2.8 | 20.3 | 9 |
|  | HL | 0.10 | [0.03, 0.34] | 2.0E-04 |  |  |  | 10 |
|  | LH | 0.62 | [0.35, 1.11] | 0.11 |  |  |  | 57 |
|  | LL | 0.74 | [0.48, 1.15] | 0.18 |  |  |  | 97 |
| CD3+ CD8+ | HH | 0.39 | [0.10, 1.61] | 0.19 | 7.5E-08 | 5.7 | 22.0 | 9 |
|  | HL | 0.12 | [0.03, 0.49] | 0.003 |  |  |  | 14 |
|  | LH | 0.67 | [0.39, 1.14] | 0.14 |  |  |  | 51 |
|  | LL | 0.85 | [0.55, 1.29] | 0.44 |  |  |  | 77 |
| CD3+ FOXP3+ | HH | 0.30 | [0.09, 1.04] | 0.06 | 2.9E-08 | 1.7 | 29.9 | 11 |
|  | HL | 0.18 | [0.08, 0.42] | 1.0E-04 |  |  |  | 16 |
|  | LH | 0.93 | [0.53, 1.60] | 0.78 |  |  |  | 34 |
|  | LL | 0.60 | [0.39, 0.92] | 0.02 |  |  |  | 80 |

\*HH=high abundance/high spatial clustering; HL = high abundance/low spatial clustering; LH = low abundance/high spatial clustering; LL = low abundance/low spatial clustering; None = samples with no positive cells is the reference group.

#### 3.3.2. Tumor compartment of TMAs

Similar to the analysis of the ROIs, the analysis of the TMAs found that EOC patients with high abundance of CD3+, CD8+, FOXP3+, and CD3+CD8+ cells and low spatial clustering had the best OS, with this difference being significant (**Figure 4B, Table 2**). The EOC patients with no abundance of any of the immune markers tended to have the worse survival. In an analysis of the subset of EOC patients with high abundance of the TILs (CD3+), the difference in survival between patients with low and high clustering of TILs was statistically significance with better survival being observed for patients with low clustering of TILs (HR = 0.16, 95% CI (0.04, 0.64)) (**Supplemental Table 2**).

### 3.4. Analysis of co-occurrence of CD3+CD8+ and CD3+FOXP3+ and ovarian cancer survival

Bivariate analysis involving Ripley’s *K* was completed to assess co-occurrence of cytotoxic T cells (CD3+CD8+) and regulatory T cells (CD3+FOXP3+), treating the CD3+FOXP3+ cell as the reference or “anchor” cell type. Results from the association of the measure of co-location with OS is presented in **Figure 5** and **Table 3**. The results using CD3+CD8+ as the reference cell were similar (data not shown). Among the ROIs, patients with low co-occurrence of cytotoxic T-cells (CD3+CD8+) and regulatory T-cells (CD3+FOXP3+) had the best survival, noting only 11 patients were in this group. In contrast, patients with high co-occurrence of cytotoxic and regulatory T cells had the worst survival (N = 166). However, these results were not confirmed in the TMA study. In the TMA analysis, patients with high co-occurrence had the best survival in the TMAs (N = 86). Additionally, when restricting analysis to samples in which both immune cell types are present in the ROIs, the EOC patients with low co-occurrence of cytotoxic T-cells and regulatory T-cells had better OS (HR = 0.463, CI = [0.26, 0.83], p-value = 0.009). However, similar analysis for the TMAs found that EOC patients with high co-occurrence of these two immune cells had better survival (HR = 1.762, CI = [1.14, 2.73], p-value = 0.0115).

**Table 3:**
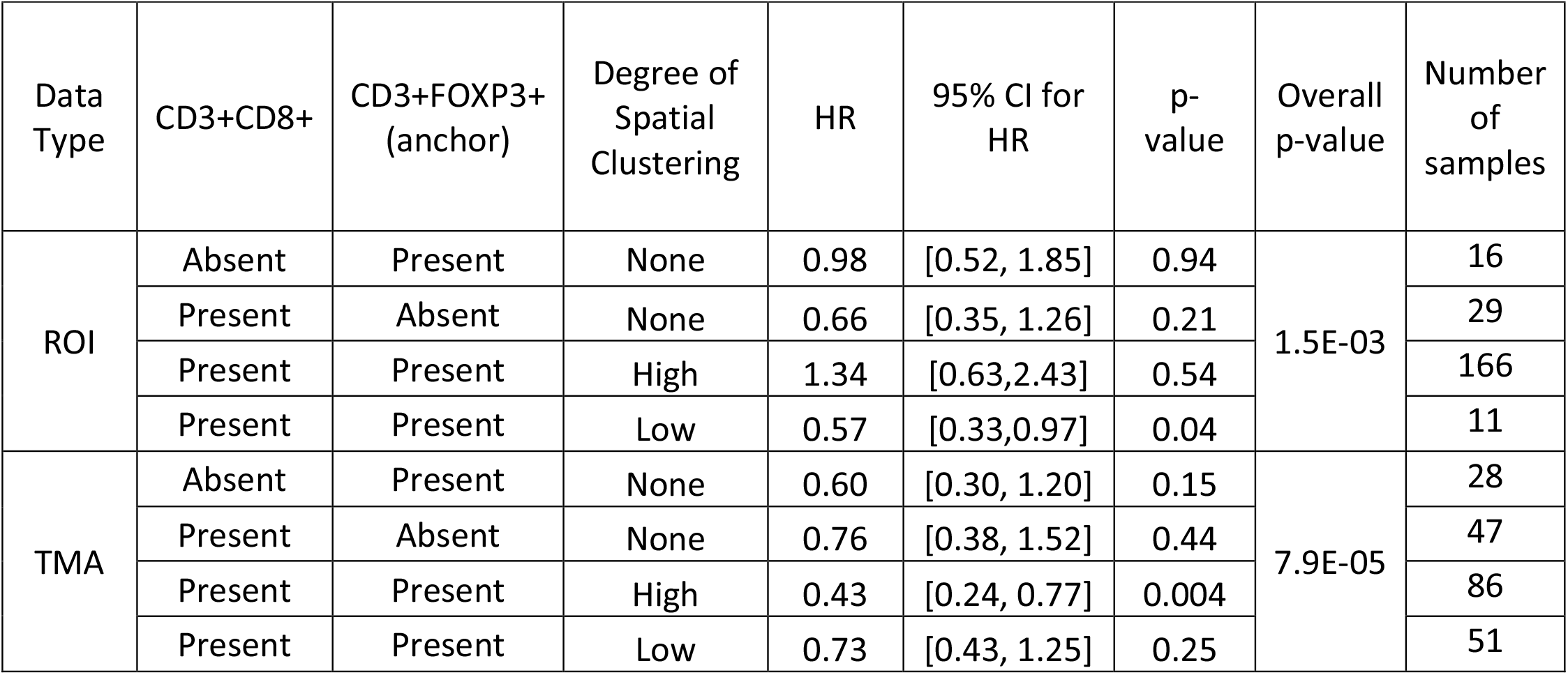
Results of co-occurrence of CD3+CD8+ (cytotoxic T-cells) and CD3+FOXP3+ (regulatory T-cells) using bivariate Ripley’s K for the ROIs and TMAs. Degree of spatial clustering based on permutation-based estimate of CSR. Models were adjusted for age at diagnosis and stage. The no cytotoxic T-cells and no regulatory T-cells (none) group/category is the reference group.

| Data Type | CD3+CD8+ | CD3+FOXP3+ (anchor) | Degree of Spatial Clustering | HR | 95% CI for HR | p-value | Overall p-value | Number of samples |
| --- | --- | --- | --- | --- | --- | --- | --- | --- |
| ROI | Absent | Present | None | 0.98 | [0.52, 1.85] | 0.94 | 1.5E-03 | 16 |
|  | Present | Absent | None | 0.66 | [0.35, 1.26] | 0.21 |  | 29 |
|  | Present | Present | High | 1.34 | [0.63, 2.43] | 0.54 |  | 166 |
|  | Present | Present | Low | 0.57 | [0.33, 0.97] | 0.04 |  | 11 |
| TMA | Absent | Present | None | 0.60 | [0.30, 1.20] | 0.15 | 7.9E-05 | 28 |
|  | Present | Absent | None | 0.76 | [0.38, 1.52] | 0.44 |  | 47 |
|  | Present | Present | High | 0.43 | [0.24, 0.77] | 0.004 |  | 86 |
|  | Present | Present | Low | 0.73 | [0.43, 1.25] | 0.25 |  | 51 |

### 3.5. Analysis of co-occurrence of CD3+CD8+ and CD3+FOXP3+ and ovarian cancer survival

Bivariate analysis involving Ripley’s *K* was completed to assess co-occurrence of cytotoxic T cells (CD3+CD8+) and regulatory T cells (CD3+FOXP3+), treating the CD3+FOXP3+ cell as the reference or “anchor” cell type. Results from the association of the measure of co-location with OS is presented in **Figure 5** and **Table 3**. The results using CD3+CD8+ as the reference cell were similar (data not shown). Among the ROIs, patients with low co-occurrence of cytotoxic T-cells (CD3+CD8+) and regulatory T-cells (CD3+FOXP3+) had the best survival, noting only 11 patients were in this group. In contrast, patients with high co-occurrence of cytotoxic and regulatory T cells had the worst survival (N = 166). However, these results were not confirmed in the TMA study. In the TMA analysis, patients with high co-occurrence had the best survival in the TMAs (N = 86). Additionally, when restricting analysis to samples in which both immune cell types are present in the ROIs, the EOC patients with low co-occurrence of cytotoxic T-cells and regulatory T-cells had better OS (HR = 0.463, CI = [0.26, 0.83], p-value = 0.009). However, similar analysis for the TMAs found that EOC patients with high co-occurrence of these two immune cells had better survival (HR = 1.762, CI = [1.14, 2.73], p-value = 0.0115).

## 4. Discussion and Conclusion

In this research, we present a novel permutation-based analysis framework using Ripley’s *K* (univariate and bivariate) to explore the relationship between the degree of spatial clustering of immune cells with a clinical outcome. The application of this framework to study of the Time of EOC tumors from 158 african American EOC patients revealed that not only the abundance of

In this research, we present a novel permutation-based analysis framework using Ripley’s *K* (univariate and bivariate) to explore the relationship between the degree of spatial clustering of immune cells with a clinical outcome. The application of this framework to study of the TIME of EOC tumors from 158 African American EOC patients revealed that not only the abundance of immune cells but also the degree of spatial clustering of immune cells within the TIME were associated with overall survival. EOC patients with high abundance and low spatial clustering of tumor-infiltrating lymphocytes (TILs) and cytotoxic T-cells had the best overall survival compared to EOC patients with tumors with no involvement of the studied immune cells. In contrast, patients with low levels and high spatial clustering of regulatory T-cells had better overall survival. These findings underscore the prognostic importance of evaluating not only immune cell abundance, but also the spatial contexture of the immune cells in the TIME.

Comparison of the value of Ripley’s *K* under the assumption of CSR (complete spatial randomness) based on theoretical derivation based on area or based on the permutation-based estimate found that the theoretical value overestimates the true value of Ripley’s *K* under CSR (**Figure 3**), with the bias more pronounced when the level of missing cells or “holes” is large (**Figure 2**). This will result in a biased estimate of the degree of spatial clustering that would impact downstream association analyses (i.e., biased and incorrect hazard ratios, confidence intervals and p-values). Many of the proposed methods being used for the spatial analysis of digital pathology data being applied, particularly in the setting of TMAs, such as nearest neighbor distances are not correcting for this “missingness” in cell measurements and thus are prone to incorrect estimation of the degree of clustering/co-localization. An additional strength of the proposed statistical framework is that the degree of clustering can be estimated for an entire TMA or ROI or focused on just the tumor or stromal compartments.

However, there are many challenges in completing the spatial analysis of the TIME, with many areas requiring further research. One challenge in using Ripley’s *K* is selection of the proximity parameter (r or radius). Often, the selection of this value is based upon prior knowledge or based on practical considerations. For the present analysis of ovarian cancer tumors, we chose to use Ripley’s *K* a r = 25 to measure the level of clustering of immune cells in a small area (or radius). A possible choice of the proximity parameter is to compute Ripley’s *K* at several values of *r* and select the value that has the greatest difference from CSR(14, 32). However, this implementation would likely lead to different proximity parameters being used for each image and would make the spatial measure not comparable across images. Another approach would be to treat the estimates of Ripley’s *K* as the various r values as a function or trajectory and applying functional data analysis (FDA), which allows for linking entire spatial trajectory to be associated with a phenotype(33-35).

Another challenge that arises when applying Ripley’s *K* is that many samples may have zero cells that express the marker of interest (i.e., “immune cold tumors”). For these cases, spatial clustering is undefined. To accommodate this case in the survival analysis, a category was defined in which samples with zero abundance and no spatial clustering was constructed. This challenge was amplified in the bivariate analysis in which both cell types had to be present in the sample for estimation of spatial co-localization. Additionally, using the optimal cut-point is a popular method for determining categories for a continuous variable (i.e., percent abundance, density), however, these methods have been shown to inflate the type I error rate (36-39) with the optimal cut-point varying between studies.

In conclusion, this paper illustrates a permutation-based approach for estimating the degree of spatial clustering when studying the TIME. This approach addresses the unique challenges of tumor samples, such as, regions where cells cannot be measured due to the limitation of the sample preparation. The application of this approach also showed that in African American patients with EOC that not only the abundance but the level of spatial clustering of CD3+ and CD3+CD8+ cells in the tumor is related to survival, where EOC patients with low level of clustering had better survival compared to those with high level of spatial clustering. The application of this spatial analysis framework to the study of the TIME could lead to the identification of immune content and spatial architecture that could aid in the determination of patients that are likely to response to immunotherapies. Additionally, the proposed spatial analysis approach is also applicable to the analysis of data off technologies that are being used to study the TIME, such as CyTOF (time-of-flight mass cytometry) and imaging mass cytometry (IMC).

## Data Availability

Access to the AACES mIF data used in testing and illustrating the method will be made available upon reasonable request to the AACES investigators, please contact Dr. Lauren Peres.

## Acknowledgments

We would like to thank the AACES investigators, Drs. Anthony Alberg, Elisa Bandera, Jill Barnholtz-Sloan, Melissa Bondy, Michele Cote, Ellen Funkhouser, Patricia Moorman, Edward Peters, Ann Schwartz, and Paul Terry, for their contributions to the AACES. We would also like to acknowledge the support of the Advanced Analytical and Digital Pathology Core under the Pathology Department at Moffitt Cancer Center.

## Author Contributions

BLF, TG, CW and LP conceived and planned the study. SMS completed the panel design, staining and the multi-spectra scanning. JN preformed the digital image analysis and quantification. CW, RT, JC, and BLF completed the statistical and bioinformatics analysis. BLF supervised the project. JS, LP provided access to the samples and data used to test the spatial analysis method. CW, RT, and JC developed the R package *spatialTIME* used in the analysis of the IF data. CW and BLF wrote the manuscript. All authors reviewed and edited the manuscript.

## Software and Data Availability Statement

The R package *spatialTIME* [under review at CRAN] was developed to implement the statistical analyses outlined in the paper and can be found at R CRAN and at [https://github.com/FridleyLab/spatialTIME]. Question about the method and R package can be submitted to. Access to the AACES mIF data used in testing and illustrating the method will be made available upon reasonable request to the AACES investigators – please contact Dr. Lauren Peres.

## Supplemental Figure Legends

**Supplemental Figure 1:** Example displaying the behavior of Besag’s L when a point process exhibits clustering, complete spatial randomness, and dispersion (left to right). The locations of the points are plotted in panel A, and the trajectory of Besag’s L - r with different values of r is plotted in panel B showing that as points are more evenly spread that Besag’s L – r decreases.

**Supplemental Figure 2:** The relationship between the percent of CD3+ cells and degree of spatial clustering has a exponentially decaying relationship (A). Plots (B) and (D) (colored red in plot A), and (C) and (E) (colored green in plot A) are two ROIs which have two approximately the same percent of CD3+ but different levels of spatial clustering.

**Supplemental Figure 3:** Histograms showing the difference between the theoretical and permutation estimate of CSR for 254 ROIs and 5 cell types. The black dashed vertical line corresponds to 0.

**Supplemental Figure 4:** Histograms showing the quantity (left column) and percentage (right column) of CD3+ cells removed from ROIs (top row) and TMAs (bottom row) due to them being classified as both CD3+CD8+ and CD3+FOXP3+.

**Supplemental Figure 5:** Scatter plots showing the cytoplasm intensity, which is used to classify cell positivity, for FOXP3 (Opal 540) and CD8 (Opal 570). These four plots show varying degree of phenotype misclassification and illustrates the challenge of making univariate or bivariate intensity threshold for classifying higher dimensional spaces.

## Supplemental Tables

**Supplemental Table 1**: Summary of the demographics of the 408 women in the AACES study, and by type of available images.

**Supplemental Table 2**: Results from survival analysis of the immune marker abundance and spatial clustering from the ROIs and TMAs only considering samples with high abundance. Degree of spatial clustering based on permutation-based estimate of CSR. Models were adjusted for age at diagnosis and stage. Group HH is reference group

## References

1. Ripley BD. Modelling Spatial Patterns. Journal of the Royal Statistical Society Series B (Methodological). 1977;39(2):172–212.

2. Ribas A, Wolchok JD. Cancer immunotherapy using checkpoint blockade. Science. 2018;359(6382):1350–5.

3. Couzin-Frankel J. Breakthrough of the year 2013. Cancer immunotherapy. Science. 2013;342(6165):1432–3.

4. Pardoll DM. The blockade of immune checkpoints in cancer immunotherapy. Nat Rev Cancer. 2012;12(4):252–64.

5. Fridman WH, Zitvogel L, Sautes-Fridman C, Kroemer G. The immune contexture in cancer prognosis and treatment. Nat Rev Clin Oncol. 2017.

6. Fridman WH, Pages F, Sautes-Fridman C, Galon J. The immune contexture in human tumours: impact on clinical outcome. Nat Rev Cancer. 2012;12(4):298–306.

7. Gooden MJ, de Bock GH, Leffers N, Daemen T, Nijman HW. The prognostic influence of tumour-infiltrating lymphocytes in cancer: a systematic review with meta-analysis. Br J Cancer. 2011;105(1):93–103.

8. Lee CW, Ren YJ, Marella M, Wang M, Hartke J, Couto SS. Multiplex immunofluorescence staining and image analysis assay for diffuse large B cell lymphoma. J Immunol Methods. 2020;478:112714.

9. Schwen LO, Andersson E, Korski K, Weiss N, Haase S, Gaire F, et al. Data-Driven Discovery of Immune Contexture Biomarkers. Frontiers in oncology. 2018;8:627.

10. Jawhar NMT. Tissue Microarray: A rapidly evolving diagnostic and research tool. Ann Saudi Med. 2009;29(2):123–7.

11. Vayrynen SA, Zhang J, Yuan C, Vayrynen JP, Dias Costa A, Williams H, et al. Composition, Spatial Characteristics, and Prognostic Significance of Myeloid Cell Infiltration in Pancreatic Cancer. Clin Cancer Res. 2021;27(4):1069–81.

12. Rose CJ, Naidoo K, Clay V, Linton K, Radford JA, Byers RJ. A statistical framework for analyzing hypothesized interactions between cells imaged using multispectral microscopy and multiple immunohistochemical markers. J Pathol Inform. 2013;4(Suppl):S4.

13. Tsakiroglou AM, Fergie M, Oguejiofor K, Linton K, Thomson D, Stern PL, et al. Spatial proximity between T and PD-L1 expressing cells as a prognostic biomarker for oropharyngeal squamous cell carcinoma. Br J Cancer. 2020;122(4):539–44.

14. Magurran AE. Biological diversity. Current biology: CB. 2005;15(4):R116–8.

15. Dunn KW, Kamocka MM, McDonald JH. A practical guide to evaluating colocalization in biological microscopy. Am J Physiol Cell Physiol. 2011;300(4):C723–42.

16. Kather JN, Suarez-Carmona M, Charoentong P, Weis C-A, Hirsch D, Bankhead P, et al. Topography of cancer-associated immune cells in human solid tumors. Elife. 2018;7:e36967.

17. Schwen LO, Andersson E, Korski K, Weiss N, Haase S, Gaire F, et al. Data-Driven Discovery of Immune Contexture Biomarkers. Front Oncol. 2018;8:627-.

18. Siegel RL, Miller KD, Jemal A. Cancer statistics, 2020. CA Cancer J Clin. 2020;70(1):7–30.

19. Schildkraut JM, Alberg AJ, Bandera EV, Barnholtz-Sloan J, Bondy M, Cote ML, et al. A multi-center population-based case-control study of ovarian cancer in African-American women: the African American Cancer Epidemiology Study (AACES). BMC Cancer. 2014;14:688.

20. Ovarian Tumor Tissue Analysis C, Goode EL, Block MS, Kalli KR, Vierkant RA, Chen W, et al. Dose-Response Association of CD8+ Tumor-Infiltrating Lymphocytes and Survival Time in High-Grade Serous Ovarian Cancer. JAMA Oncol. 2017;3(12):e173290.

21. Peres LC, Cushing-Haugen KL, Kobel M, Harris HR, Berchuck A, Rossing MA, et al. Invasive Epithelial Ovarian Cancer Survival by Histotype and Disease Stage. Journal of the National Cancer Institute. 2019;111(1):60–8.

22. Besag, J.E.. Discussion on Dr Ripley’s Paper. Journal of the Royal Statistical Society: Series B (Methodological). 1977;39(2):192–212.

23. Baddeley A, Rubak E, Turner R. Spatial point patterns: methodology and applications with R. Boca Raton; London; New York: CRC Press, Taylor & Francis Group; 2016. xvii, 810 pages p.

24. Gabriel E.A. Baddeley, E. Rubak, R. Turner: Spatial Point Patterns: Methodology and Applications with R. Mathematical Geosciences. 2017;49(6):815–7.

25. Perry GLW, Miller BP, Enright NJ. A comparison of methods for the statistical analysis of spatial point patterns in plant ecology. Plant Ecology. 2006;187(1):59–82.

26. Moore M. Spatial Statistics: Methodological Aspects and Applications. New York, NY: Springer-Verlag; 2001. 282 p.

27. Cressie N, Ver Hoef JM. Spatial statistical analysis of environmental and ecological data.404-9.

28. Good P. Springer Series in Statistics. 1994.

29. Bouwmeester W, Zuithoff NP, Mallett S, Geerlings MI, Vergouwe Y, Steyerberg EW, et al. Reporting and methods in clinical prediction research: a systematic review. PLoS medicine. 2012;9(5):1–12.

30. Mabikwa OV, Greenwood DC, Baxter PD, Fleming SJ. Assessing the reporting of categorised quantitative variables in observational epidemiological studies. BMC health services research. 2017;17(1):201.

31. Altman DG, Lausen B, Sauerbrei W, Schumacher M. Dangers of using “optimal” cutpoints in the evaluation of prognostic factors. Journal of the National Cancer Institute. 1994;86(11):829–35.

32. Kiskowski MA, Hancock JF, Kenworthy AK. On the use of Ripley’s K-function and its derivatives to analyze domain size. Biophysical journal. 2009;97(4):1095–103.

33. Cardot H, Ferraty F, Sarda P. Functional Linear Model. Statistics & Probability Letters. 1999;45(1):11–22.

34. Ramsay JO, Dalzell CJ. Some Tools for Functional Data Analysis. Journal of the Royal Statistical Society: Series B (Methodological). 1991;53(3):539–61.

35. Ramsay JO, Silverman BW. Applied functional data analysis: methods and case studies. New York: Springer; 2002.

36. Hilsenbeck SG, Clark GM. Practical p-value adjustment for optimally selected cutpoints. Statistics in medicine. 1996;15(1):103–12.

37. Lausen B, Schumacher M. Maximally Selected Rank Statistics. Biometrics. 1992;48(1):73–85.

38. Lausen B, Schumacher M. Evaluating the effect of optimized cutoff values in the assessment of prognostic factors. Computational Statistics & Data Analysis. 1996;21(3):307–26.

39. Miller R, Siegmund D. Maximally Selected Chi Square Statistics. Biometrics. 1982;38(4):1011–6.

