## Supplementary figures and images for "Statistical framework for studying the spatial architecture of the tumor immune microenvironment"

### Supplementary Figure 1

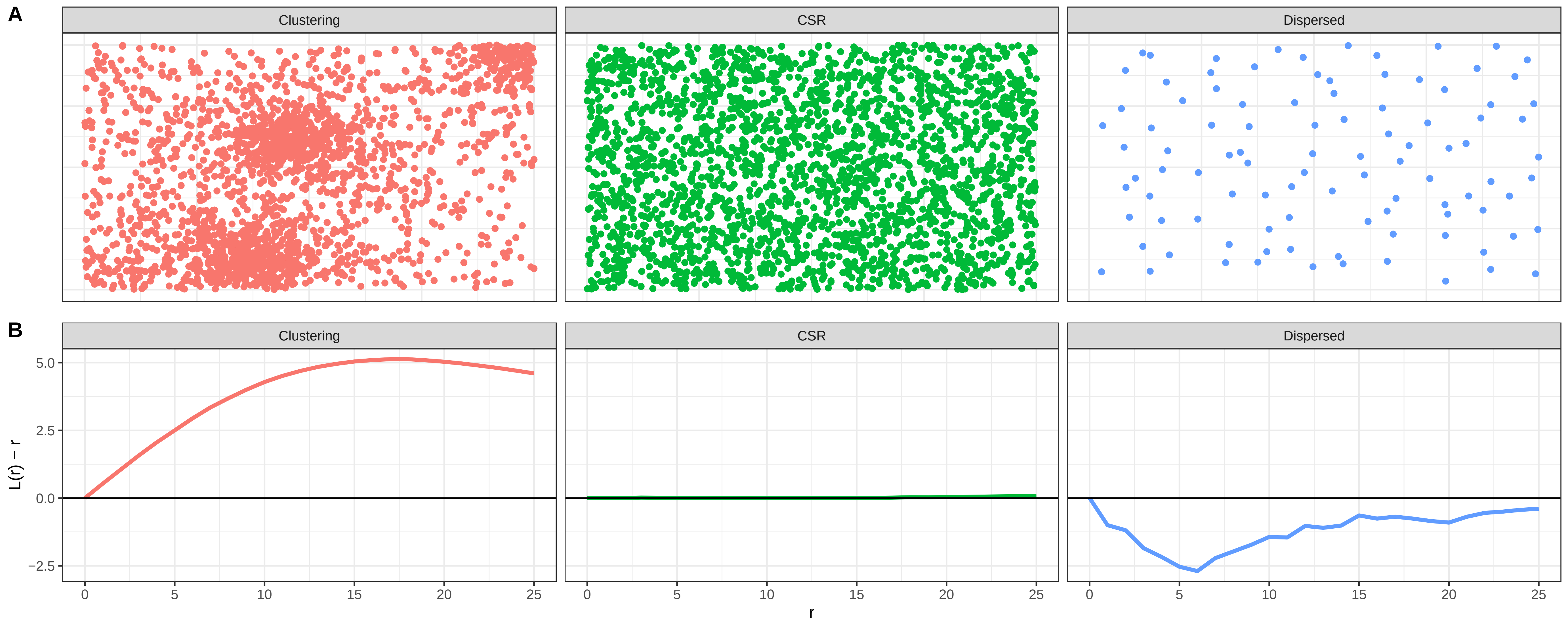

### Supplementary Figure 2

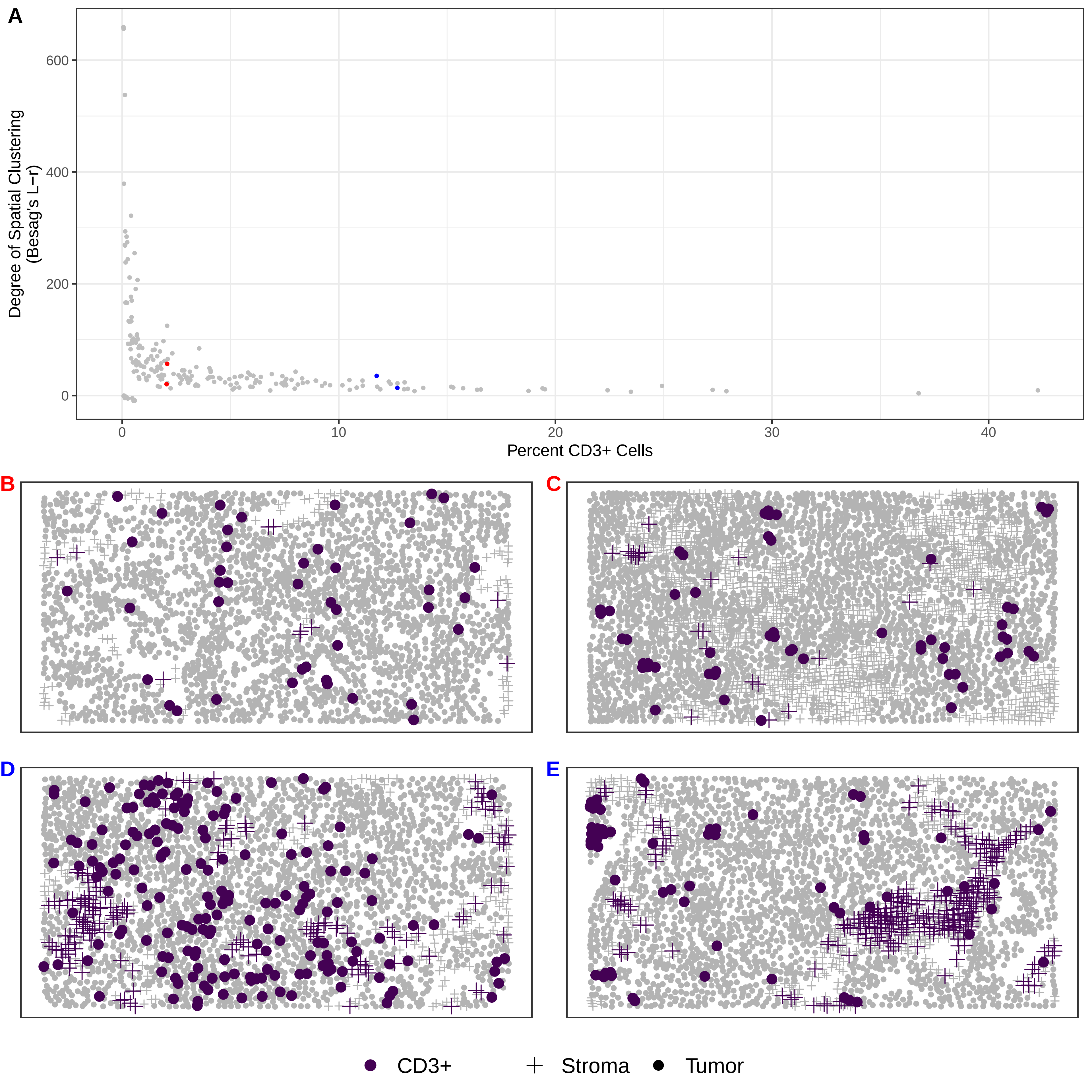

### Supplementary Figure 3

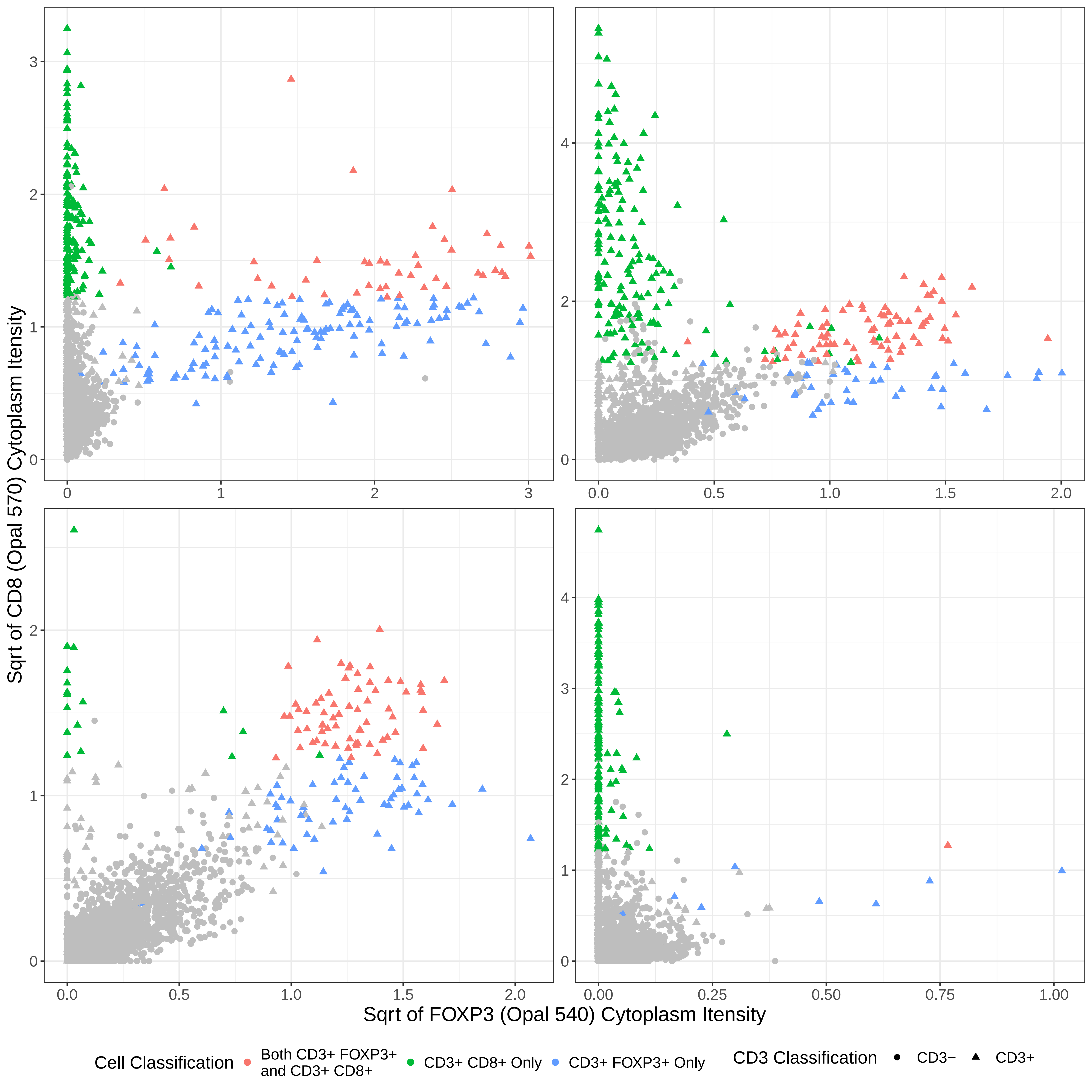

### Supplementary Figure 4

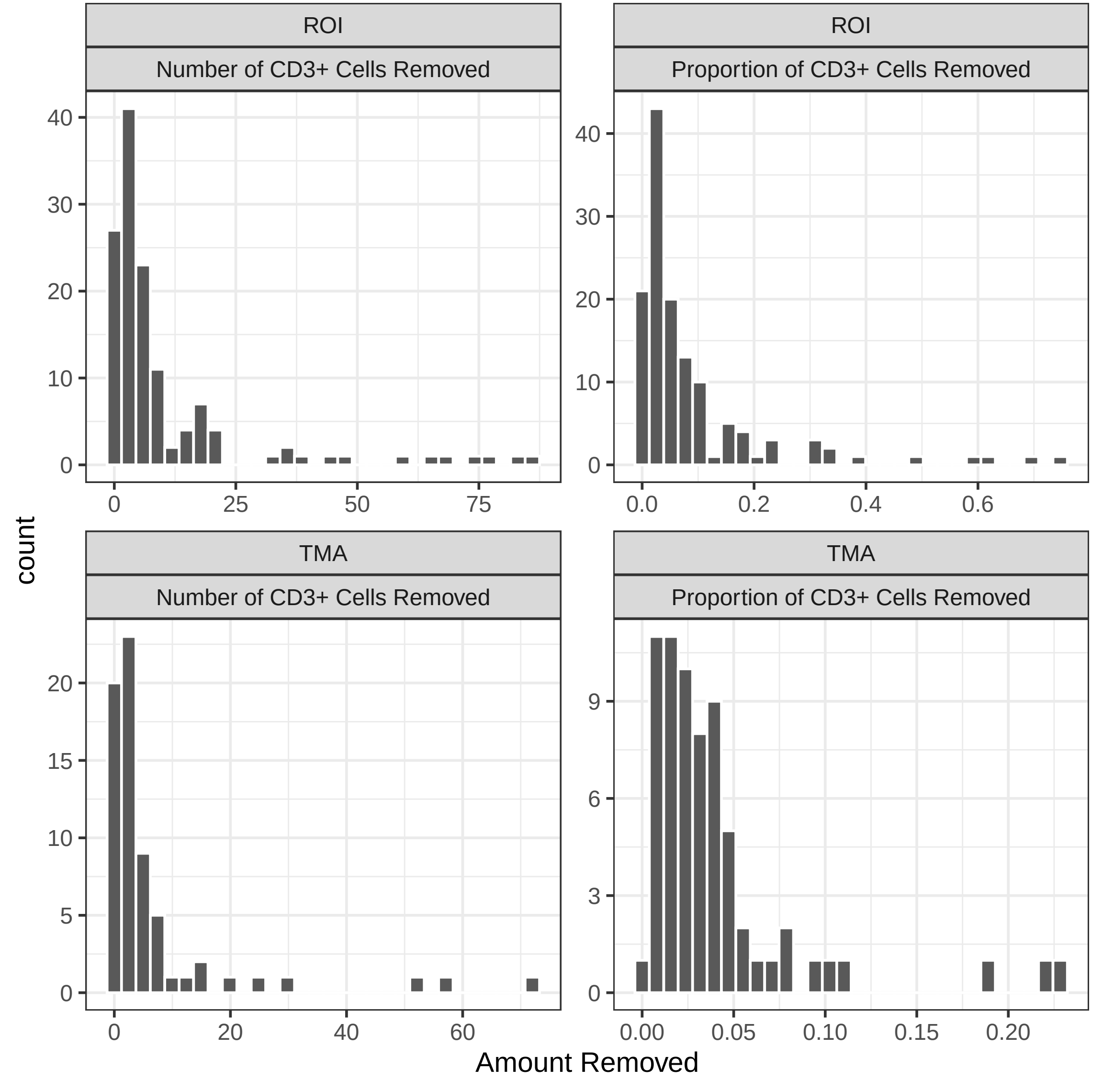

### Supplementary Figure 5

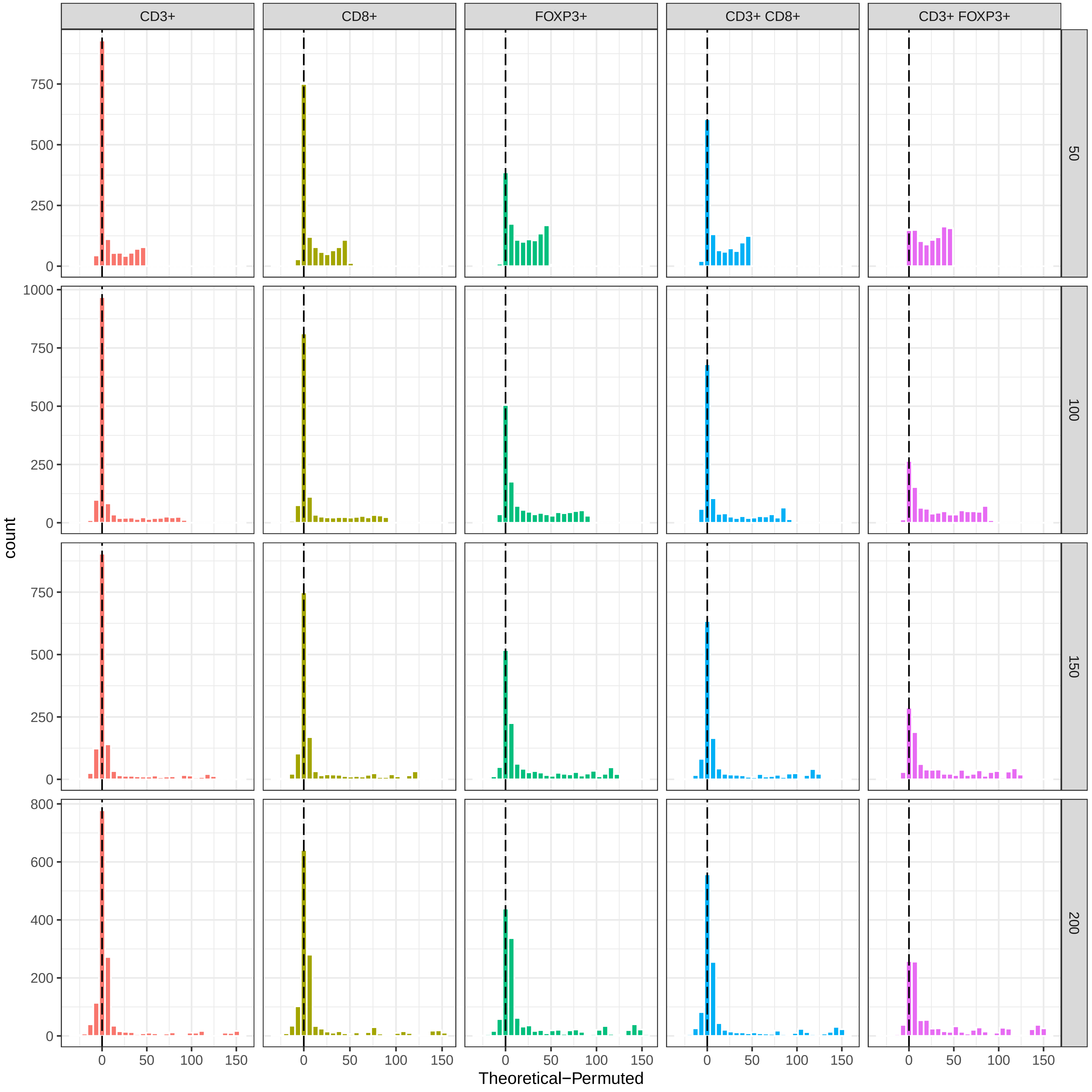
